# Effect of thiazolidinediones compared to other antidiabetic medications on incident dementia in people with type 2 diabetes: A target trial emulation study

**DOI:** 10.64898/2026.03.18.26348742

**Authors:** Bright C. Ofoha, Yuan Tian, Li-Chia Chen, Martin K. Rutter, Eleni Domzaridou, Dorina Cadar, David A. Jenkins, Matthew Sperrin, Joyce Yun-Ting Huang

## Abstract

**Background:** People with Type 2 diabetes mellitus (T2DM) are at increased risk of developing dementia. Evidence suggests that thiazolidinediones (TZDs) may be protective for dementia onset including Alzheimer’s disease and vascular dementia, compared to other second-line antidiabetic medications (SAMs). However, causality remains uncertain due to methodological limitations. We examined the effect of TZD on the risk of vascular dementia and all-cause dementia in T2DM, compared to other second-line treatments.

**Methods:** We emulated a pragmatic randomised trial using UK primary care data, Clinical Practice Research Datalink Aurum, between 2003 and 2023 to estimate the comparative effectiveness of initiating a TZD, dipeptidyl peptidase-4 (DPP-4) inhibitors, sodium-glucose cotransporter-2 (SGLT2) inhibitors, or sulfonylurea (SU) against incident dementia in T2DM adults on metformin therapy. Patients were followed for up to 5 years from 180 days after their first SAM prescription. We used overlap weighting to adjust for baseline confounding and fitted double robust Cox models to estimate adjusted hazard ratios (aHRs).

**Findings:** This study included 124,311 participants (mean age 63 years, 61% males, and 20% whites), of whom 595 developed vascular dementia and 1,678 developed all-cause dementia during follow-up. On top of metformin, 8,669 initiated TZD, 30,216 initiated DPP-4 inhibitors, 55,997 initiated SU and 29,429 initiated SGLT2 inhibitors. TZD were associated with a similar risk of vascular dementia compared with DPP-4 inhibitors (aHR 0.89;95% CI 0.36–2.23) and SU (0.58;0.24–1.42). SGLT2 inhibitors were associated with a lower risk of vascular dementia than TZD (0.29;0.09–0.94), DPP-4 inhibitors (0.25;0.10–0.64), and SU (0.17;0.07–0.40). Most patterns persisted in all-cause dementia: SGLT2 inhibitors vs DPP-4 inhibitors (0.51;0.26– 0.99) and SGLT2 inhibitors vs SU (0.35;0.18–0.67), with no difference observed between SGLT2 inhibitors and TZDs.

**Interpretation:** Dementia risk was similar for TZDs, DPP-4inhibitors and SUs but was significantly lower for SGLT2 inhibitors, a finding that warrants further investigation. Considering potential cognitive effects when selecting therapies for T2DM is important in an ageing population.

**Funding:** This work was funded by Alzheimer’s Research UK (grant: ARUK-PPG2023B-036) and the National Institute for Health and Care Research (NIHR) Greater Manchester Patient Safety Research Collaboration (GM PSRC).

**Research in context:** *Evidence before this study:* We searched for relevant studies in PubMed database with the search term (“thiazolidinedione” OR “TZD” OR “pioglitazone” OR “rosiglitazone”) AND (“diabet*” OR “T2D” OR “T2DM”) AND (“dementia*” OR “cognitive impairment” OR “Alzheimer*”) from inception until June 15, 2025. We identified 12 relevant cohort studies that examined the association between thiazolidinedione (TZD) and vascular dementia, as well as Alzheimer’s disease and all-cause dementia. The evidence from these studies is inconsistent; two of the studies suggested that TZD use was associated with a significant reduction in risk of vascular dementia, while one suggested that the risk was comparable. Current evidence suggests that TZD use were associated with a reduced risk of incident Alzheimer’s disease and all-cause dementia. Preclinical studies suggest that TZDs may reduce the risk of vascular dementia by improving cerebral perfusion, enhancing endothelial function, and reducing impairments in learning, memory, and blood–brain barrier permeability. However, these findings are limited using a non-representative sample, insufficient adjustment for confounding, inadequate outcomes definition, and an increased risk of a false-positive result purely by chance.

*Added value of this study:* To our knowledge, this is the first study in the UK to compare the effectiveness of TZDs with other second-line antidiabetic medications on the risk of vascular and all-cause dementia. To address methodological limitations in prior studies, we applied a rigorous causal inference framework and emulated a pragmatic randomised trial using a large, nationally representative primary care electronic health record dataset to minimise bias. We found that initiating TZDs as second-line therapy provided a comparable risk of vascular dementia and all-cause dementia as other second-line medications. Sodium-glucose cotransporter-2 (SGLT2) inhibitors were associated with a lower risk of vascular dementia than TZDs, and with a reduced risk of both vascular dementia and all-cause dementia compared with dipeptidyl peptidase-4 inhibitors and sulfonylureas.

*Implications of all the available evidence:* Our findings do not support a cognitive benefit of TZDs when used as a second-line therapy for type 2 diabetes mellitus. In contrast, SGLT2 inhibitors may offer cognitive benefits, complementing their recognised cardiovascular and renal benefits, although further research is required to confirm their causal effects.

## Introduction

Dementia prevention has been identified as a global public health priority,^1^ and evidence-based policies are urged to reduce dementia risk. More than 55 million people currently live with dementia worldwide, with around 10 million new cases occur each year; a figure projected to triple by 2050.^2^ In 2024, the global economic burden of dementia was estimated at US $1.3 trillion.^2^ Vascular dementia is the second most common dementia subtype, affecting around 180,000 people in the UK,^3^ which costs society £42 billion annually.^4^ Alzheimer’s disease (AD) accounts for up to 80% of the UK’s 982,000 dementia cases, making it the most common dementia subtype.^5^ However, unlike AD, there is no approved treatment for vascular dementia.

Vascular dementia results from reduced cerebral blood flow caused by damaged or obstructed blood vessels, leading to progressive cognitive impairment, including memory loss, confusion, inattention, and language difficulties.^6,7^ These vascular injuries cause permanent neurological damage. People with vascular dementia have a shorter life expectancy of about five years, compared with eight to ten years for people with AD, largely due to a higher risk of stroke, myocardial infarction, and other vascular events.^8^ Managing underlying vascular conditions, such as hypertension, stroke, diabetes, and atherosclerosis, may help lower the incidence of vascular dementia.^3^ AD, in contrast, arises from distinct pathological mechanism, involving the abnormal accumulation of amyloid-β and tau proteins in the brain. These accumulations form pathological deposits known as amyloid plagues and neurofibrillary tangles, which disrupt the normal neuronal function.^9^ Recently licenced antibody treatments for early-stage AD, such as lecanemab and donanemab, target and slow down the progression of amyloid plaques; however, these treatments are currently unavailable in the NHS due to their high cost.^10,11^

People with type 2 diabetes mellitus (T2DM) have an increased risk of both vascular dementia and AD, primarily driven by chronic hyperglycaemia and insulin resistance, which damage cerebral vessels, accelerate amyloid accumulation, and promote neuroinflammation.^12,13^ Thiazolidinediones (TZDs), a class of oral antihyperglycemic agents that activate peroxisome proliferator-activated receptors to improve insulin sensitivity, have shown potential neuroprotective effects.^14–16^ Animal experimentation suggest that TZDs may reduce cerebral vascular injury and improve endothelial function, learning and memory, and blood–brain barrier integrity.^17,18^ Prior observational studies suggested that TZD use is associated with a lower risk for incident vascular dementia, AD and all-cause dementia in people with T2DM, ^15,16,19–25^ while other observational studies suggested otherwise. ^14,26,27^

Several methodological limitations in these studies make it challenging to draw a conclusion on the potential neuroprotective effect of TZD, such as using a non-representative sample, short study time frame which limits follow-up (≤ 5 years), increased risk of false positive results, and insufficient adjustment of measured confounding with standard regression or fewer covariates (4 to 5 variables).^16,19–28^ In addition, missing data may not have been handled adequately, leaving the possibility of residual confounding.^16,19–28^ To date, no randomised controlled trials have directly compared the relationship between these antidiabetic medications and dementia outcomes. More importantly, UK-based evidence on the potential neuroprotective effects of TZD use against dementia remains scarce, highlighting the need for further research.

This study aimed to assess the comparative effectiveness of second-line TZDs versus dipeptidyl peptidase-4 (DPP-4) inhibitors, sodium-glucose cotransporter-2 (SGLT2) inhibitors, and sulfonylureas (SUs) on the risk of incident vascular dementia and all-cause dementia among people with T2DM, treated with metformin. We applied a pragmatic target trial emulation within a causal inference framework to minimise common sources of bias in pharmacoepidemiological research. These selected four drug classes are routine oral antidiabetic drugs, commonly prescribed at a similar stage of T2DM condition. When first-line treatment fails to control patient’s glycated haemoglobin (HbA1c) beyond an agreed threshold any of these second-line treatment could be prescribed for a patient, in accordance with the National Institute for Health and Care Excellence (NICE) guidance on managing T2DM. Therefore, our hypothetical pragmatic randomised trial compares the relative effectiveness of these four second-line drug classes with direct relevance to real-world prescribing practice.

## Methods

### Study design and data sources

This retrospective cohort study, emulating a pragmatic target trial, used UK primary care electronic health records from the Clinical Practice Research Datalink (CPRD) Aurum database, from January 2003 to December 2023. CPRD contains anonymised UK primary care records that are broadly representative of the UK population and was linked to the Office for National Statistics Death Registry and the patient-level Index of Multiple Deprivation (IMD) data. However, only general practices in England primarily contribute data to CPRD Aurum currently.^29^ The study protocol was approved by the CPRD Independent Scientific Advisory Committee on 14 August 2024 (Protocol number: 24_003967).

We emulated the protocol of a pragmatic target trial to estimate the comparative effectiveness of initiating TZDs, DPP-4 inhibitors, SGLT2 inhibitors, or SUs on a background of metformin therapy, on risk for incident dementia. The target trial estimated the treatment effectiveness of being assigned to initiate TZD versus DPP-4 inhibitor, SGLT2 inhibitor, or SU therapy, added to metformin. Key elements of the target trial protocol, including the eligibility criteria, treatment strategies being compared, assignment procedures, follow-up period, outcomes of interest, causal contrasts of interest, and the analysis plan, are summarised in Supplementary **Table S1**. Reporting adhered to the TARGET (TrAnsparent ReportinG of studies Emulating a Target trial) guidelines,^30^ with a completed checklist in Supplementary **Table S2**.

### Eligibility criteria

Eligible participants were adults (≥18 years) with: (i) a recorded diagnosis of T2DM in CPRD Aurum during the study period; (ii) ≥1 year of prior registration with a GP practices in England that has agreed to contribute data to CPRD before treatment escalation; and (iii) successfully linked to ONS mortality and IMD data (iv) prescription for metformin monotherapy prior to initiation of a second-line antidiabetic medication (SAM), and ≥1 metformin prescription within 6 months after initiation of a SAM. We excluded individuals with any non-T2DM diabetes type and those with prior dementia or cognitive impairment.

We also excluded individuals initiating second-line agents outside the four strategies under comparison. The study schematic and timeline are presented in **Figure 1**.

**Figure 1.**
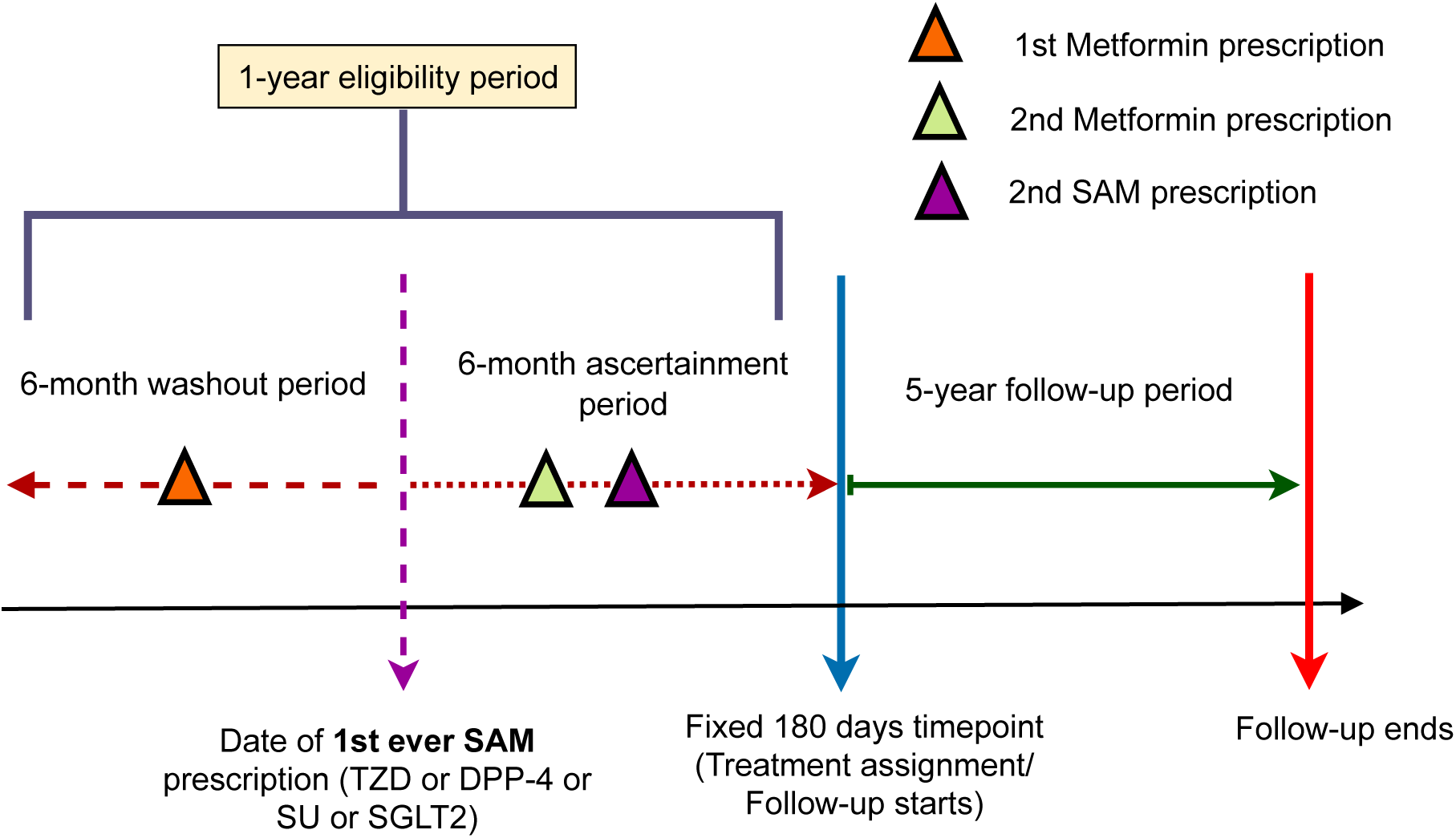
**Study design and timelines** This timeline illustrates the identification of new and stable users of second-line antidiabetic medications (SAMs) added to metformin, their assignment to treatment strategies, and follow-up from 1 January 2003 to 31 December 2023. DPP-4 = dipeptidyl peptidase-4 inhibitors; SGLT2 = sodium-glucose cotransporter 2 inhibitors; SU = sulfonylureas; TZD = thiazolidinediones

### Treatment strategies and emulated randomisation

At treatment escalation, the strategies corresponded to initiating one of four SAMs (TZDs, DPP-4 inhibitors, SGLT2 inhibitors or SUs) added to metformin. Treatment initiation was defined as the first prescription of the SAM added to metformin, confirmed by a second prescription of the same SAM within 180 days to improve the specificity of treatment initiation and reduce early-exposure misclassification. The first SAM prescription date was defined as the index date which corresponds as the cohort entry date. Medication exposures were identified using CPRD Product Codes, which maps to the Dictionary of Medicines and Devices, a subset of SNOMED CT. For brevity, we refer to the groups as TZD, DPP-4 inhibitors, SGLT2 inhibitors, and SU (each added to metformin).

Although treatment allocation in the observational data was not random, we emulated random assignment by overlap weighting of generalised propensity scores (GPS) estimated from baseline characteristics measured prior to treatment initiation. Overlap weighting targets the average treatment effect in the overlap population and creates a pseudo-population with improved covariate balance across treatment groups, mimicking attributes of a pragmatic randomised trial.^31,32^

### Outcomes and follow-up period

The primary outcome was incident vascular dementia diagnosed during follow-up, identified using Medication Codes, which primarily map to SNOMED CT. A new vascular dementia case was defined as the first recorded diagnosis of vascular dementia, either as the first dementia diagnosis in a participant’s record or following a prior diagnosis of another dementia subtype. In alignment with established research practices to enhance diagnostic validity, we defined confirmed cases as individuals with at least two vascular dementia codes recorded at least 30 days apart,^14,15^ with the first diagnosis recorded as the event date. The secondary outcome was all-cause dementia, defined and validated using the same approach.

To align eligibility, treatment assignment, and start of follow-up, we used 180-day landmark after the first SAM prescription as time zero. Follow-up then continued for up to five years, or until transfer out of practice, dementia diagnosis, death, or the study end (31 December 2023), whichever occurred first. This landmark approach ensured consistent alignment of eligibility, treatment assignment, and start of follow-up across all participants.^33^ By defining eligibility, treatment status, start of follow-up at the landmark time, the landmark design helps prevent immortal time bias arising from delayed treatment assignment.^34^ We chose 180-day landmark time to align with NICE recommendation; that HbA1c level should be measured every 90 to 180 days, HbA1c level is the key factor that informs treatment regimen. ^35^ We excluded individuals that died or had dementia before the landmark time.

### Covariates and data handling

Twenty-three potential confounders were prespecified based on clinical relevance to treatment choice and dementia risk (Supplementary **Table S3**), including sociodemographic characteristics (age, sex, ethnicity, and IMD quintile), comorbidities (stroke, chronic kidney disease, hypertension, heart disease, diabetic microvascular complications, and peripheral vascular disease), concurrent medications use (anticoagulants, antiplatelets, statins, antihypertensives, anticholinergics, antidepressants, and antipsychotics), lifestyle factors (body mass index (BMI) and smoking status), and biomarkers (HbA1c and systolic blood pressure (SBP)).

Sociodemographic characteristics, comorbidities, and lifestyle factors were ascertained from records obtained within 1 year prior to SAM initiation; concurrent medications and biomarkers were captured within 180 days prior to SAM initiation. For multiple measurements recorded on the same date, we used the mean value. We defined BMI, smoking status, HbA1c, and SBP using published algorithms (Supplementary **File S1**). ^36–38^

We quantified the proportion of missing data for each covariate. Under the assumption that missingness can be fully explained by the observed data (missing at random), we used multiple imputation by chained equations (MICE) to impute missing baseline values, including HbA1c, SBP, BMI, IMD quintile, and smoking status. Given the high level of missingness in ethnicity data (>70%), records with missing values were combined with those labelled “not recorded” to form a single category. Five imputed datasets were created, and all analyses were repeated in each dataset; estimates were then pooled using Rubin’s rules to account for imputation uncertainty.

### Statistical analyses

We estimated intention-to-treat pairwise treatment effects for all six comparisons (TZD vs DPP-4 inhibitors, TZD vs SGLT2 inhibitors, TZD vs SU, SGLT2 inhibitors vs DPP-4 inhibitors, SGLT2 inhibitors vs SU, and DPP-4 inhibitors vs SU) between the four SAMs using double-robust Cox models with overlap weighting. To induce the attributes of random treatment allocation of the target trial, we adjusted for baseline confounding using the overlap weighting – a propensity score weighting method. Overlap weighting estimates the average treatment effect in the overlap population, defined as patients with similar baseline characteristics across treatment groups. Given the moderate to poor covariate overlap across treatment groups, as shown by the distributions of the GPS across treatment groups (Supplementary **Figure S1**), overlap weighting was applied to focus inference on the region of common support.^39^ GPS were defined as the probability of receiving each specific SAM of interest, conditional on observed patient baseline characteristics. GPS were estimated using multinomial logistic regression, including all prespecified twenty-three baseline covariates. All continuous variables were modelled using restricted cubic splines with five knots to avoid imposing an unverifiable linearity assumption (Supplementary **Table S3**). Two interaction terms (age × BMI and SBP × HbA1c) were included in the GPS model to account for their joint influence on treatment selection, which improved covariate balance.

Double-robust Cox models were used to estimate adjusted hazard ratios (aHRs) for each pairwise treatment comparison. These models adjusted for all baseline confounders included in the GPS model, excluding interaction terms. Robust standard errors were used to account for within-patient correlation arising from the overlap weighting procedure. The double-robust approach helps mitigate residual confounding arising from possible misspecification of the GPS model, thereby improving statistical efficiency. To control the family-wise error rate across the six pairwise comparisons, we applied Tukey’s multiple-comparison correction.^40^ Proportional hazards assumptions were evaluated using Schoenfeld residual tests (each covariate had p > 0.05) and visual inspection of residual plots. Statistical analyses were performed using the R statistical programme (version 4.4.2).

We performed four sensitivity analyses to assess the robustness of our findings:

i. Competing risk analysis: To account for death as a competing risk, we estimated subdistribution hazard ratio (sHR) using Fine–Gray model.
ii. Alternative exposure definition: Treatment initiation was redefined as either ≥2 prescriptions of the same SAM within 90 days, observed twice in 6-month intervals, or ≥2 prescriptions within one year, with gaps >90 and <180 days. The definition of metformin monotherapy use remained the same, which is a first prescription within 180 days prior to SAM initiation, and a second prescription within 6 months following SAM initiation.
iii. Complete case analysis: Standard Cox proportional hazard models were fitted using only participants with complete data for all covariates.
iv. Long follow-up period: We did not restrict the follow-up to 5 years. Therefore, follow-up began 180 days after the first SAM prescription and ended when a participant was transferred out of practice, diagnosed with dementia, died, or the study ended (31 December 2023), whichever occurred first.

## Results

A total of 124,311 adults with T2DM were included in this study. Of these, 8,669 (7%) initiated a TZD, 30,216 (24%) a DPP-4 inhibitor, 55,997 (45%) an SU, and 29,429 (24%) an SGLT2 inhibitor as second-line therapy. The cohort selection is summarised in **Figure 2**, and baseline characteristics of the study population are presented in **Table 1**. Participants initiating SGLT2 inhibitors were slightly younger (mean age 60 years) than those in the TZD, SU, or DPP-4 inhibitors group (mean age 62–64 years).

**Figure 2.**
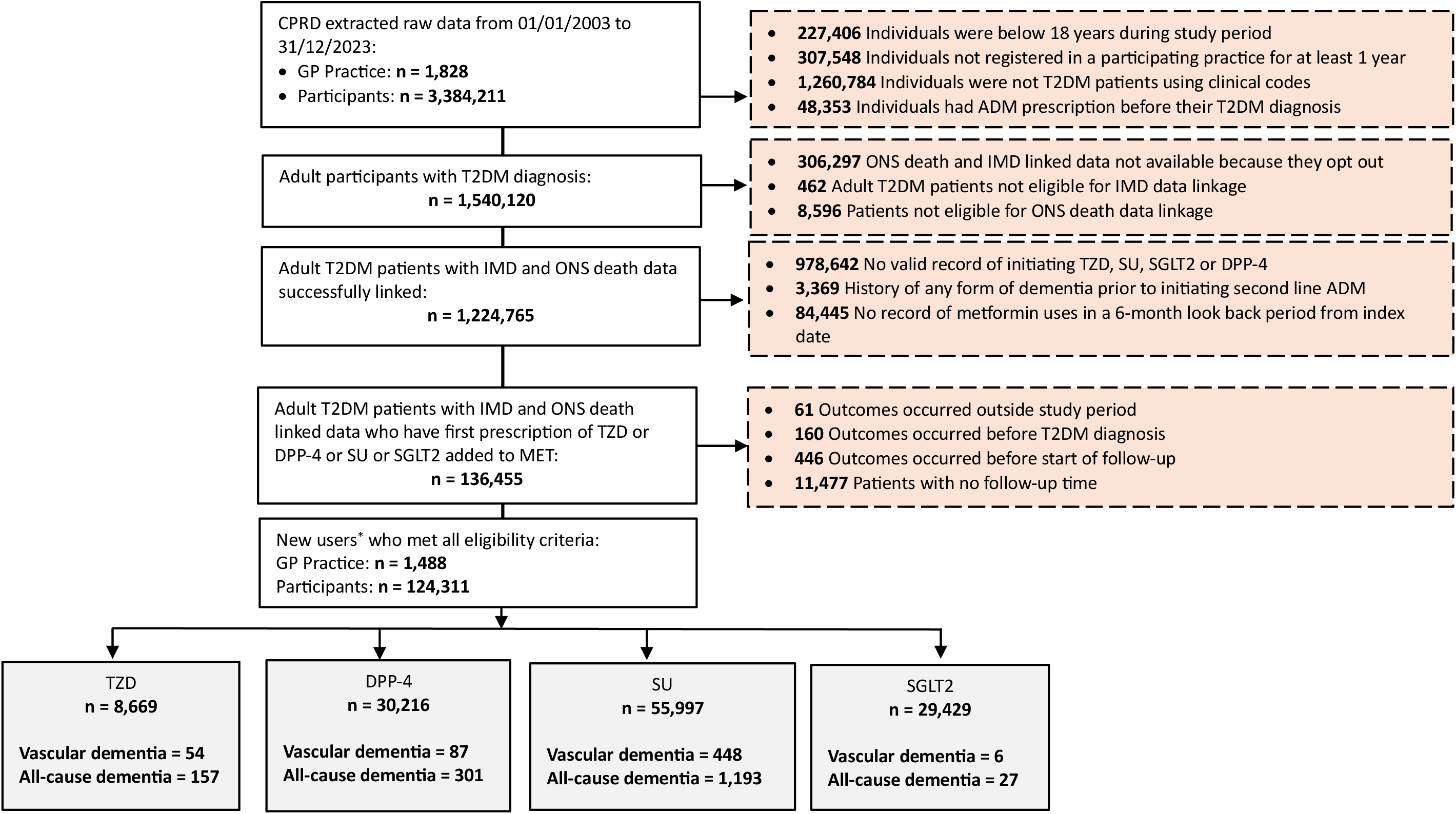
**Cohort selection flowchart** *The follow-up of eligible individuals started with successfully linkage to ONS mortality and IMD data of those who met data quality metric (i.e. acceptable quality for research) and ended up with the earliest time of transfer out dates, death dates, dementia diagnosis dates, last collection dates, end of study dates and event dates. Abbreviations: CPRD: Clinical Practice Research Datalink; GP: General Practice; T2DM: Type 2 Diabetes Mellitus; ADM: Antidiabetic Medication; ONS: Office for National Statistics; IMD: Index of Multiple Deprivation; MET: Metformin; TZD: thiazolidinedione; DPP-4: dipeptidyl peptidase-4 inhibitor; SGLT2: sodium–glucose cotransporter-2 inhibitor; SU: sulfonylurea.

**Table 1.**
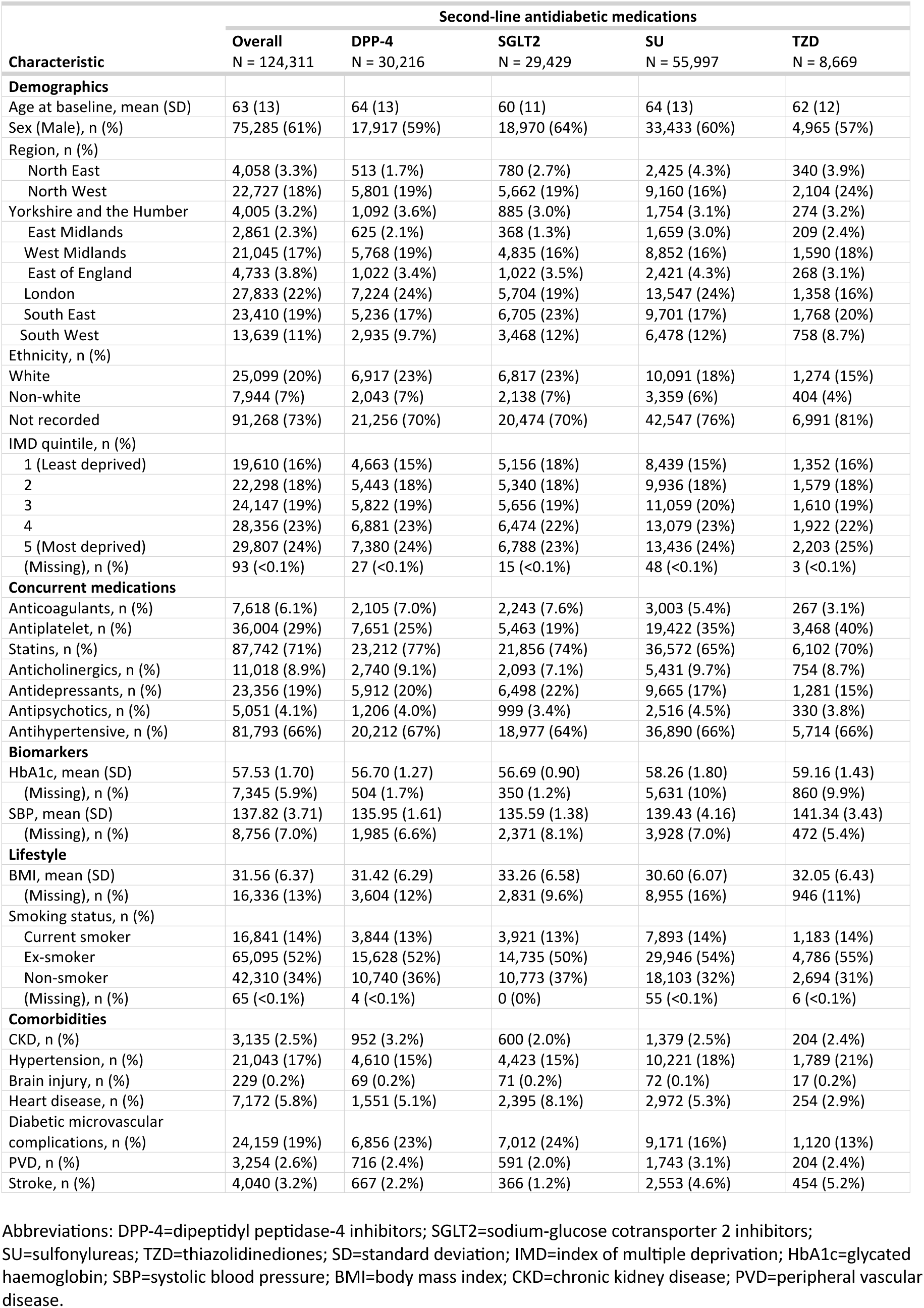
Baseline characteristics of the study population, stratified by second-line antidiabetic medications.

The SGLT2 inhibitors group also had a higher proportion of males (64%) than the SU (60%), DPP-4 inhibitors (59%), or TZD (57%) groups. There was no notable difference in IMD quintile distribution across treatment groups.

People in the SGLT2 inhibitors group showed the highest proportion of anticoagulant use (7.6%) and heart disease (8.1%) among the treatment groups but had the lowest percentage of stroke (1.2%). In contrast, people in the TZD group demonstrated the highest percentage of stroke (5.2%), accompanied by the highest proportion of antiplatelet use (40%). Diabetic microvascular complications occurred more frequently in the SGLT2 inhibitors and DPP-4 inhibitors groups (23–24%) than in the SU and TZD groups (13–16%). People in the SU and TZD groups tended to have higher HbA1c levels (58–59 mmol/mol) than those in the DPP-4 inhibitor and SGLT2 inhibitor groups (56–57 mmol/mol). All treatment groups had a mean BMI>30.

To assess if the baseline covariates were balanced across the treatment groups after weighting, we calculated the standardised mean differences (SMD) before and after weighting (Supplementary **Table S4)**. After applying overlap weighting, all covariates had an SMD < 0.1, indicating that the covariates were balanced across treatment groups.

During the 5-year follow-up, the SGLT2 inhibitors group had the lowest incidence rate of vascular dementia (11 cases per 100,000 person-years) compared to DPP-4 inhibitors, TZD and SU (93–230 cases per 100,000 person-years) (Supplementary **Table S5**). **Figure 3** presents the pairwise comparisons of incident vascular dementia risk between treatment groups. TZD use was not significantly associated with vascular dementia risk compared with DPP-4 inhibitors (aHR 0.87; 95% CI 0.36–2.27) and SUs (aHR 0.57; 95% CI 0.23–1.47).

**Figure 3.**
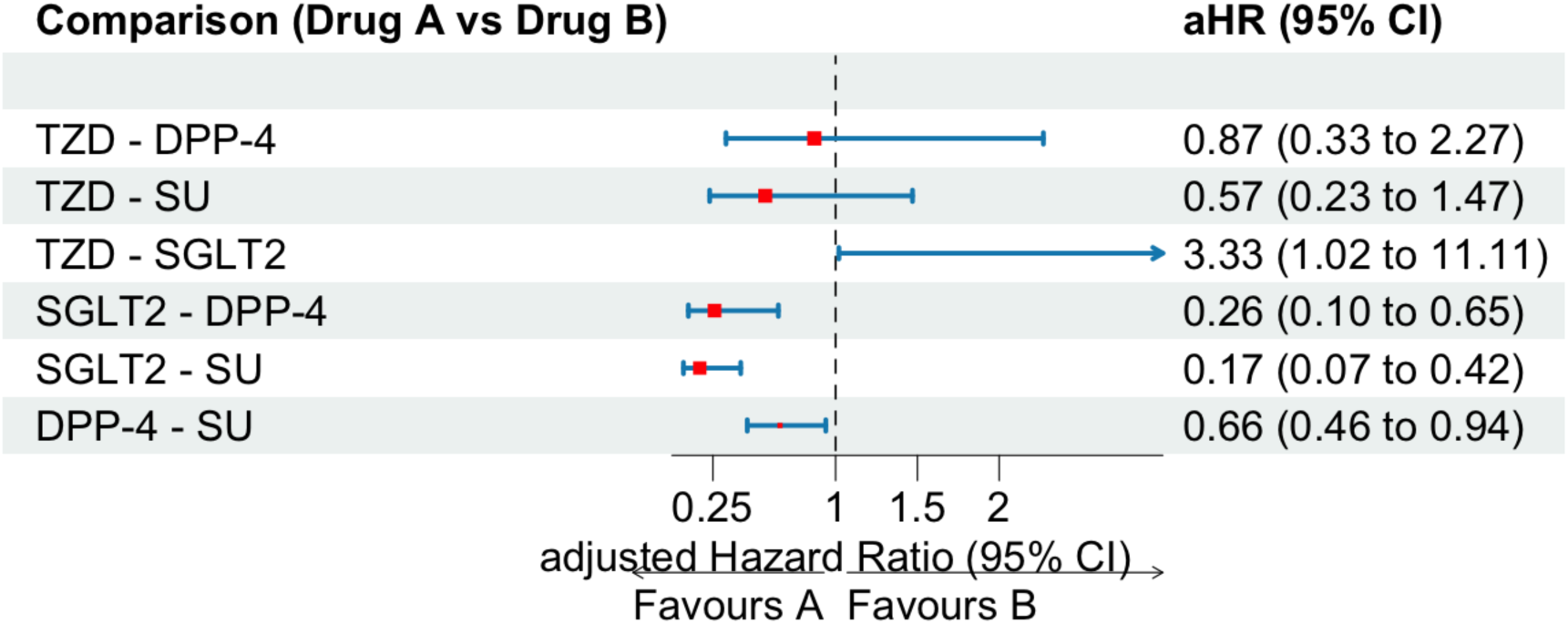
**Pairwise comparisons of incident vascular dementia risk between second-line antidiabetic medications** Adjustments for multiple comparisons was done using Tukey’s method. Models were adjusted for age at baseline, sex, region, ethnicity, Index of Multiple Deprivation, comorbid stroke, chronic kidney disease, hypertension, heart disease, diabetic microvascular complications, peripheral vascular disease, concurrent use of anticoagulants, antiplatelets, statins, antihypertensives, anticholinergics, antidepressants, and antipsychotics, as well as biomarkers (systolic blood pressure, HbA1c) and lifestyle (body mass index, smoking status). Abbreviations: HR = hazard ratio; CI = confidence interval; TZD = thiazolidinedione; DPP-4 = dipeptidyl peptidase-4 inhibitor; SGLT2 = sodium–glucose cotransporter-2 inhibitor; SU = sulfonylurea.

Compared with SGLT2 inhibitors, TZD was associated with a higher risk of vascular dementia and estimates were imprecise (aHR 3.33; 95% CI 1.02–11.11). SGLT2 inhibitors were associated with a lower risk of vascular dementia compared with DPP-4 inhibitors (aHR 0.26; 95% CI 0.10–0.65) and SUs (aHR 0.17; 95% CI 0.07–0.42). Compared with SUs, initiation of DPP-4 inhibitors was associated with a lower risk of vascular dementia (aHR 0.66; 95% CI 0.46–0.94).

For all-cause dementia, SGLT2 inhibitors also had the lowest incidence rate (51 cases per 100,000 person-years) compared to DPP-4 inhibitors, TZD and SU (321–612 cases per 100,000 person-years) (Supplementary **Table S5**). The results of the comparative effectiveness of SAMs on all-cause dementia were similar to those for vascular dementia (**Figure 4**). TZD was not significantly associated with all-cause dementia risk compared with DPP-4 inhibitors (aHR 0.94; 95% CI 0.53–1.67) and SU (aHR 0.64; 95% CI 0.37–1.12).

**Figure 4.**
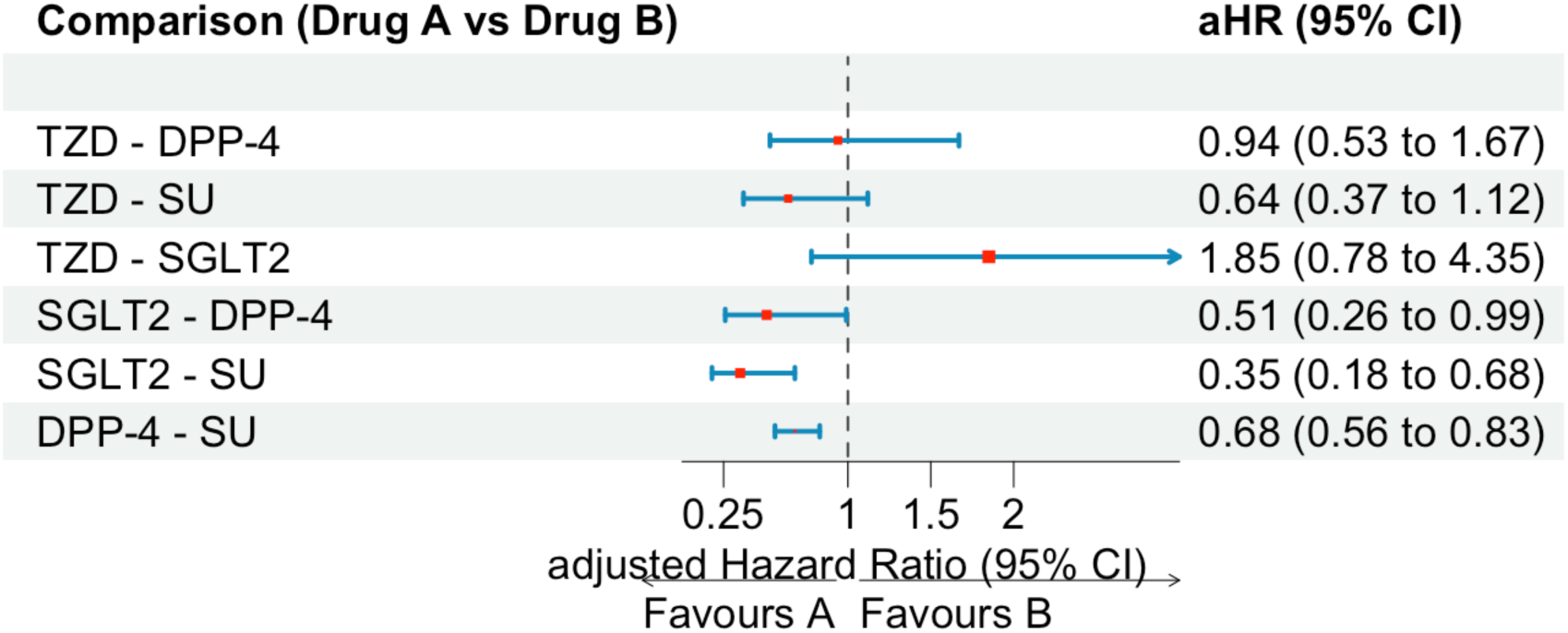
**Pairwise comparisons of incident all-cause dementia risk between second-line antidiabetic medications** Adjustments for multiple comparisons was done using Tukey’s method. Models were adjusted for age at baseline, sex, region, ethnicity, Index of Multiple Deprivation, comorbid stroke, chronic kidney disease, hypertension, heart disease, diabetic microvascular complications, peripheral vascular disease, concurrent use of anticoagulants, antiplatelets, statins, antihypertensives, anticholinergics, antidepressants, and antipsychotics, as well as biomarkers (systolic blood pressure, HbA1c) and lifestyle (body mass index, smoking status). Abbreviations: HR = hazard ratio; CI = confidence interval; TZD = thiazolidinedione; DPP-4 = dipeptidyl peptidase-4 inhibitor; SGLT2 = sodium–glucose cotransporter-2 inhibitors; SU = sulfonylurea.

Compared with SGLT2 inhibitors, TZD was not significantly associated with all-cause dementia risk (aHR 1.85; 95% CI 0.78–4.35). As with vascular dementia, SGLT2 inhibitors were associated with a lower risk of all-cause dementia compared with DPP-4 inhibitors (aHR 0.51; 95% CI 0.26–0.99) and SU (aHR 0.35; 95% CI 0.18–0.68). Compared with SU, initiation of DPP-4 inhibitors was associated with a lower risk of all-cause dementia (aHR 0.68; 95% CI 0.56–0.83).

Results from sensitivity analyses were generally consistent across comparisons involving SU and DPP-4 inhibitors, but estimates comparing SGLT2 inhibitors and TZD were sensitive to initiation definitions and follow-up assumptions. The competing risk analyses that accounted for death yielded results similar to the primary findings, confirming the robustness of this study (Supplementary **Table S6**). When exposure was defined using stricter criteria, initiators of TZDs were not significantly associated with vascular dementia risk compared to those initiating SGLT2 inhibitors (aHR 3.45; 95% CI 0.93–12.5), but a higher risk of all-cause dementia with imprecise estimates (aHR 2.86; 95% CI 1.27–6.67) (Supplementary **Table S7**).

In complete case analysis, SGLT2 inhibitors were not significantly associated with vascular dementia risk, compared to TZD (aHR 2.94; 95% CI 0.93–9.09) (Supplementary **Table S8**). Similarly, when the follow-up period was not restricted to 5 years, there was no significant difference in vascular dementia risk between TZD users and SGLT2 inhibitor users (aHR 3.03; 95% CI 0.89–10.00) (Supplementary **Table S9**).

## Discussion

This study found that people who initiated TZDs as second-line treatment showed a comparable risk of vascular dementia and all-cause dementia compared with other second-line medications. Results suggest that SGLT2 inhibitors might be beneficial than TZDs in preventing vascular dementia but not all-cause dementia. The use of SGLT2 inhibitors was also associated with a lower risk of both vascular dementia and all-cause dementia, compared with initiating DPP-4 inhibitors and SUs.

Despite promising results with SGLT2 inhibitors, their potential benefits on cognition warrant further investigation, particularly given the small number of vascular dementia events observed in the SGLT2 inhibitor group and their known cardioprotective effects. The discrepancy between findings for vascular dementia and all-cause dementia when comparing SGLT2 inhibitors with TZDs may be partly explained by the differing number of events observed for the primary and secondary outcomes.

The primary findings were partly consistent across sensitivity analyses. The potential protective effect of SGLT2 inhibitors against vascular dementia compared with TZDs was observed in the competing risk analysis but not in analyses with stricter exposure definition, people with complete record only, or extended follow-up. These findings suggest that the observed association may not be robust across alternative analytical assumptions, therefore should be interpreted cautiously.

Sensitivity analyses using a stricter exposure definition and complete-case analysis substantially reduced the sample size, thereby lowering statistical power and limiting the ability to detect a true difference, even if one existed. The results from the extended follow-up analysis may reflect several methodological issues associated with longer follow-up, including higher loss to follow-up, greater impact of time-varying confounding, treatment misclassification, and survival bias (where individuals remaining at later follow-up are generally healthier). These factors may attenuate the observed effect of SGLT2 inhibitors.

### Comparison with existing literature

Most prior observational studies have reported a lower risk of dementia among people with T2DM treated with TZDs, when compared to other therapies.^16,19–25,28^ However, we did not observe such an association for either vascular or all-cause dementia. Direct comparison with previous studies is challenging because our study assessed the add-on effect of second-line therapies to metformin, whereas most studies compared monotherapy regimens (e.g., TZDs vs. metformin). Although previous studies, including one from Korea and two from Taiwan, have also examined dual therapy and reported cognitive benefits associated with TZD use,^16,19,22^ this was not replicated in our study. A possible explanation is our use of a target trial emulation, which applied strict eligibility criteria and achieved better balance in baseline characteristics across treatment groups. This rigorous study design and adjustment for multiple hypotheses testing provide more robust estimates of treatment effects and reduce the risk of false-positive findings.

Our findings suggesting potential cognitive benefits of SGLT2 inhibitors among people with T2DM are consistent with existing evidence.^41,42^ A recent multicentre cohort study using data from Korean population found that SGLT2 inhibitors were associated with reduced dementia risk including AD, compared with TZDs.^26^ Results from a network meta-analysis of observational studies showed that SGLT2 inhibitors ranked highest in cognitive benefits and were associated with a lower risk of all-cause dementia and vascular dementia compared with non-users, metformin, DPP-4 inhibitors and SU.^41^ Similarly, a meta-analysis of cohort studies found that SGLT2 inhibitors were associated with a lower risk of all-cause dementia, but not vascular dementia, compared with DPP-4 inhibitors.^42^ Additionally, a study using TriNetX data reported neuroprotective effects across all dementia subtypes among people who initiated SGLT2 inhibitors as first-line therapy compared with metformin.^43^ Although the number of dementia cases in the SGLT2 inhibitor group was small in our study (may be because they are newer and have less follow-up), the consistency of our results with prior research supports the validity and external reliability of our findings. More importantly, our study contributes new UK-specific evidence to this growing literature.

### Strengths and limitations

This study has several notable strengths. First, we used a large, nationally representative primary care electronic health records dataset spanning more than 20 years, which enabled robust estimation of dementia risk across multiple SAMs in UK general practices. Second, we used strict new-user design which included people who were stable new users of SAMs, confirmed by a second prescription of the same SAM within six months of initiation and no prior use of other antidiabetic drugs. This approach reduced the likelihood of treatment misclassification and supports an intention-to-treat design by minimising early treatment switching. Third, is the improved alignment of eligibility, treatment assignment, and the start of follow-up at a common time point (180 days after the first prescription), thereby improving internal validity and minimising biases from immortal time.^33^ With the landmark method, follow-up began at a 180-day landmark after the first second-line prescription and treatment initiation was confirmed by a repeat prescription within this window. This approach reduces early exposure misclassification and limits bias from rapid switching but may also introduce selection bias if survival during the landmark period differs across drug classes. Consequently, estimates may partly reflect a healthier or more adherent subset of initiators, particularly for comparisons involving newer therapies. We therefore interpret findings as comparative effectiveness among stable initiators. Fourth, we employed an advanced alternative propensity score weighting method, overlap weighting, to address baseline differences in biomarkers and clinical characteristics (all SMDs <0.1), when inverse probability treatment weighting (IPTW) failed to achieve acceptable balance for HbA1c and SBP (SMDs >0.1) (Supplementary **Figure S2**). This method also improves efficiency by focusing on participants with the most comparable baseline characteristics across treatment groups, thereby reducing the undue influence of extreme propensity scores.^31^ Our analytic decisions adhered closely to the principles of pragmatic target trial emulation. Finally, validated outcome definitions, correction for multiple testing, appropriate handling of missing data, and sensitivity analyses strengthen the robustness of our findings.

Our study has some limitations that should be acknowledged. As with observational studies, residual confounding from unmeasured or imperfectly measured factors (e.g., frailty, medication adherence, or functional status) cannot be completely excluded. Also, the relatively small number of vascular dementia events, particularly in the SGLT2 inhibitors group, may have reduced statistical power and precision in some comparisons. Furthermore, consistently with other studies using CPRD, drug exposure was defined based on prescriptions rather than dispensation, and medication adherence over time were not captured. Diagnostic misclassification of dementia subtype is also possible because our study relied on clinical diagnoses coded in the primary care system, and coding accuracy may vary across practices or clinicians. Lastly, ethnicity was poorly recorded (>70% missing), limiting residual confounding control.

### Implications for clinical practice

This study provides important evidence with direct implications for clinical decision-making in the management of T2DM. Our findings highlight that adding SGLT2 inhibitors as second-line therapy may offer additional benefits in reducing dementia risk, complementing their recognised cardiovascular and renal advantages.^35^ TZDs, in contrast, do not provide additional cognitive benefits for people with T2DM, beyond other conventional second-line therapies. As dementia prevention becomes an increasingly important public health priority, health professionals could proactively consider, long-term neurological effects of treatment choices, alongside glycaemic control, when managing T2DM.

The management of T2DM is also evolving. According to updated NICE guidance in February 2026, SGLT2 inhibitors are now recommended as a first-line treatment alongside metformin.^44^ Their cardiovascular and kidney protective effects are well established, but their potential cognitive benefits remain unclear.

Further investigations are needed to confirm whether SGLT2 inhibitors can help protect brain health. This includes causal analyses that consider dynamic treatment strategies and pilot clinical trials, particularly because the number of dementia cases in this study was relatively small. Future studies should also clarify the underlying mechanisms linking antidiabetic medications to brain health and examine whether treatment duration, dosage, timing from diabetes onset, or specific patient subgroups influence dementia risk.

## Conclusion

This study highlights that adding TZDs as second-line therapy had a comparable risk of vascular dementia and all-cause dementia compared with other second-line medications in T2DM. However, the addition of SGLT2 inhibitors may offer additional benefits in reducing the risk of dementia. Given the risk of dementia in people with diabetes, the long-term cognitive effects of treatment options should be considered in T2DM management.

## Contributors

BO and JYTH conceived the study. BO led the data preparation and analysis and drafted the initial manuscript. JYTH, DAJ, DC, MKR, and LCC secured funding. JYTH, DAJ, and MS supervised the study. ED helped extract CPRD data. YT helped with code lists generation. All authors contributed to the interpretation of the findings, critically reviewed the manuscript and contributed to revisions. All authors have read and approved the final manuscript.

## Ethics approval

The study was approved by the CPRD’s Independent Scientific Advisory Committee (approval number: 24_003967). CPRD also has ethical approval from the Health Research Authority to support research using anonymised patient data (research ethics committee reference 21/EM/0265). Individual patient consent was not required, as all data were deidentified.

## Data sharing

This study used pseudonymised patient-level data from the CPRD. To protect patient confidentiality, we cannot publish patient-level data. Other researchers can use patient-level CPRD data in a secure environment by submitting an application to the CPRD Independent Scientific Advisory Committee. Details of the application process and access conditions are provided by the CPRD at https://www.cprd.com/Data-access.

## Acknowledgments

This study is based on data from the Clinical Practice Research Datalink obtained under licence from the UK Medicines and Healthcare Products Regulatory Agency. The data are provided by patients and collected by the National Health Service (NHS) as part of its care and support. The Office for National Statistics mortality data are subject to Crown copyright (2026) protection and are reused with the permission of The Health and Social Care Information Centre, all rights reserved. The interpretation and conclusions contained in this study are those of the authors alone, and not necessarily those of the NIHR, NHS, or Department of Health and Social Care. We would like to acknowledge all the data providers and general practices who make anonymised data available for research.

