## Supplementary for "Effect of thiazolidinediones compared to other antidiabetic medications on incident dementia in people with type 2 diabetes: A target trial emulation study"

**Table S7 Pairwise comparisons of incident dementia risks between second-line antidiabetic medications with alternative definition of treatment initiation† (N = 88,427) 11**

**Table S1 Specification and emulation of the major components of the target trial protocol**

| Component | Pragmatic target trial | Emulator<br>Cohort analysis |
|---|---|---|
| Eligibility criteria | • Aged ≥ 18 years between 01 January 2003 and 31 December 2023<br>• Registered with a participating practice for at least a year<br>• Diagnosed with T2DM and no history of other types of diabetes including type 1<br>• No prior use of SAM in the past six months<br>• No prior dementia or other cognitive impairment causing conditions<br>• Must have at least one prescription of metformin within 6 months prior to initiating a SAM | Same with the target trial. We will identify T2DM diagnosis using CPRD Aurum SNOMED Codes |
| Treatment strategies | Initiation of any of the following SAM (added to metformin monotherapy) at baseline:<br>(a) Initiate TZD<br>(b) Initiate DPP-4 Inhibitors<br>(c) Initiate SU<br>(d) Initiate SGLT2 inhibitors | Analogues using observational data.<br>To minimise treatment switches and identify stable users of a SAM (added to metformin), one of these patterns must be established during the six months ascertainment period after the initiation of any SAM of interest at baseline:<br>• First ever prescription of SAM between 01 January 2003 and 31 December 2023<br>• A second prescription within six months of initiating a SAM |
| Treatment assignment | Eligible individuals are randomly assigned to a strategy at baseline (and must initiate treatment within 3 months), them and their healthcare workers are aware of the assigned treatment strategy | Eligible individuals are assigned to a treatment strategy before a landmark time point, based on the treatment pattern observed during ascertainment period |
| Time zero/baseline | Date when all eligibility criteria are met, and second-line treatment is initiated | 180 days (a fixed landmark time point) after all eligibility criteria are met, and a SAM is initiated |
| Follow-up period | Starts at baseline and ends on the date of the outcome, transfer out from practice, dementia diagnosis, death, 5 years after baseline or the study ends, whichever occurs first | Starts at the landmark time point and ends on the date of the outcome, transfer out from practice, dementia diagnosis, death, 5 years after follow-up starts, or the study ends, whichever occurs first |
| Outcomes | Incidence of vascular dementia and all-cause dementia (including Alzheimer's disease) during follow-up | Same as the target trial<br>We identified the outcomes using SNOMED Codes. A new case of vascular dementia will be defined as people who have a diagnosis of vascular dementia, which is the first ever diagnosis of any dementia in their records, or people who are diagnosed with vascular dementia after an initial diagnosis of dementia with a six-month diagnosis gap. A confirmed vascular dementia case is defined as two diagnosis codes that are at least 30 days apart and the first diagnosis is considered the index date |
| Causal contrast of interest | Intention-to-treat effect (ITT) | Same as the target trial |
| Statistical analysis plan | The ITT effect will be estimated by comparing the outcomes of individuals who initiated their assigned treatment (added to metformin) within a grace period of 3 months | An analogue of the target ITT effect is estimated by comparing the outcomes of the individuals who initiated and stayed on a SAM (added to metformin) for at least 6 months |

**Abbreviations**: T2DM: Type 2 diabetes mellitus; CPRD: Clinical practice research datalink; SNOMED: Systematized nomenclature of medicine–clinical terms; SAM: Second-line antidiabetic medications; TZD: Thiazolidinediones; DPP-4: dipeptidyl peptidase-4 inhibitors; SU: Sulfonylureas; SGLT2: sodium–glucose cotransporter-2 inhibitors; ITT: Intention-to-treat.

Table S2 Completed TARGET checklist of items and their location in the study report

| Item no. | | Checklist item | Location reported | |
|---|---|---|---|---|
| **Abstract** | | | | |
| **1** | a | Identify that the study attempts to emulate a target trial using observational data. State the study objectives and briefly summarize the specified target trial. | Page ii | |
| | b | Report the data sources used for emulation. | Page ii | |
| | c | Summarize key assumptions, statistical methods, findings and conclusions. | Pages ii – iii | |
| **Introduction** | | | | |
| **2** | Background | Describe the scientific background of the study and the gap in knowledge. | Pages 1 – 3 | |
| **3** | Causal question | Summarize the causal question. | Page 4 | |
| **4** | Rationale | Describe the rationale for emulating a target trial with the available data. Cite randomized trials informing the design of the target trial if applicable. | Page 4 | |
| **Methods** | | | | |
| **5** | Data sources | Cite the data sources contributing to the analyses and for each one describe the following: original purpose, type, the geographic locations, setting and time-period. If relevant, describe how the data were linked or pooled. | Page 5 | |
| **6** | **Target trial specification**<br>Specify the components of the target trial protocol that would answer the causal question. | **7** **Target trial emulation**<br>Describe how the components of the target trial protocol were emulated with the observational data, including how all variables were measured or ascertained. | Location item 6 (specification) reported | Location item 7 (emulation) reported |

| | | | | | |
|---|---|---|---|---|---|
| **Eligibility criteria** | | **Eligibility criteria** | | Page 5; Supplementary Table S1 | Page 5; Supplementary Table S1 |
| a | Describe the eligibility criteria. | a | Describe how the eligibility criteria were operationalized with the data. | | |
| **Treatment strategies** | | **Treatment strategies** | | Page 7; Supplementary Table S1 | Page 7; Supplementary Table S1 |
| b | Describe the treatment strategies that would be compared. | b | Describe how the treatment strategies were operationalized with the data. | | |
| **Assignment procedures** | | **Assignment procedures** | | Page 7; Supplementary Table S1 | Page 7; Supplementary Table S1 |
| c | Report that eligible individuals would be randomly assigned to treatment strategies and may be aware of their treatment allocation. | c | Describe how assignment to treatment strategies was operationalized with the data. | | |
| **Follow-up** | | **Follow-up** | | Page 7; Supplementary Table S1 | Page 7; Supplementary Table S1 |
| d | Clarify that follow-up would start at time of assignment to the treatment strategies. Specify when follow-up would end. | d | Clarify that follow-up starts at the time individuals were assigned to the treatment strategies. Describe how the end of follow-up was operationalized with the data. | | |
| **Outcomes** | | **Outcomes** | | Page 7; Supplementary Table S1 | Page 7; Supplementary Table S1 |
| e | Describe the outcomes. | e | Describe how the outcomes were operationalized with the data. | | |
| **Causal contrasts** | | **Causal contrasts** | | Page 9; Supplementary Table S1 | Page 9; Supplementary Table S1 |
| f | Describe the causal contrasts of interest, including effect measures. | f | Describe how the causal contrasts were operationalized with the data, including effect measures. | | |
| **Identifying assumptions** | | **Identifying assumptions** | | Page 9; Supplementary Table S1 & File S1 | Page 9; Supplementary Table S1 & File S1 |
| g | Describe assumptions that would be made to identify each causal estimand. Describe the variables, if any, related to these assumptions. | g.i | For each causal estimand, describe assumptions made to identify it, including assumptions regarding baseline confounding due to lack of randomization. | | |

| | | | | | | |
|---|---|---|---|---|---|---|
| | | | g.ii | Describe how the variables related to these assumptions were operationalized with the data | | Page 8; Supplementary File S1 |
| **Data analysis plan** | | | **Data analysis plan** | | | |
| h | For each causal estimand, describe the data analysis procedures and any associated statistical modelling assumptions, including approaches for handling missing data. | | h.i | For each causal estimand, describe the data analysis procedures and any associated statistical modelling assumptions, including approaches for handling missing data. | Pages 8-9; Supplementary Table S1 | Pages 8-9; Supplementary Table S1 |
| | | | h.ii | For each causal estimand, describe any additional analyses conducted to assess the sensitivity of the results to the choice of operationalizations, assumptions and analysis. | | |
| **Results** | | | | | | |
| **8** | Participant selection | Report numbers of individuals assessed for eligibility, eligible, and assigned to each treatment strategy. A flow diagram is strongly recommended. | | | Page 11 | |
| **9** | Baseline data | Describe the distribution of characteristics of individuals at baseline, by treatment strategy. | | | Page 12 | |
| **10** | Follow-up | Summarize length of follow-up and describe reasons for end of follow-up for each treatment strategy and causal contrast. | | | Page 13 | |
| **11** | Missing data | Describe the frequency of missing data in all variables, by treatment strategy when applicable. | | | Page 12 | |
| **12** | Outcomes | Describe the frequency or distribution of each outcome, by treatment strategy. | | | Page 11 | |
| **13** | Effect estimates | Report the effect estimates for each causal contrast with corresponding measures of precision, including both absolute and relative measures of effect, when applicable. | | | Pages 13 – 14; Figures 3 & 4 | |
| **14** | Additional analyses | Report results of all analyses to assess the sensitivity of the estimates to choices in operationalizations, assumptions and analysis. | | | Page 14 | |
| **Discussion** | | | | | | |

| | | | |
|---|---|---|---|
| **15** | Interpretation | Provide an interpretation of the key findings. | Page 16 |
| **16** | Limitations | Discuss the limitations of the study considering differences between the target trial and its emulation and the plausibility of assumptions, including assumptions regarding baseline confounding due to lack of randomization. | Page 18 |
| **Other information** | | | |
| **17** | Ethics | Provide the institutional research board or ethics committee that approved the study and approval numbers, if relevant. | Page 20 |
| **18** | Registration | State whether, when and where the study protocol was registered. | NA |
| **19** | Sharing of study materials | Provide information on whether data, analytic code and/or other materials are accessible, and where and how they can be accessed. | 21 |
| **20** | Funding sources | Provide the sources of funding and detail the role of the funders in the design, conduct and reporting of the study. | 21 |
| **21** | Conflicts of interest | State any conflicts of interest and financial disclosures for all authors. | 20 |

Observational Studies Emulating a Target Trial: The TARGET Statement. JAMA. 2025; DOI: 10.1001/jama.2025.13350
© 2025 Cashin et al. This is an Open Access article distributed under the terms of the Creative Commons Attribution-NoDerivatives License (CC BY-ND 4.0), which permits redistribution, commercial and non-commercial, provided the work is passed along unchanged and in whole, with credit to the original author(s).

### Table S3 Details of baseline covariates adjusted for in the double robust Cox models†

| S/NO | Covariates | Extraction start point | Extraction end point | Unit of measurement/Categories |
|---|---|---|---|---|
| **1** | Age at baseline | 1 year before cohort entry | Cohort entry date | Years |
| **2** | Sex | 1 year before cohort entry | Cohort entry date | 1=Male; 2=Female |
| **3** | Region | 1 year before cohort entry | Cohort entry date | 1=North East; 2=North West; 3=Yorkshire & The Humber; 4=East Midlands; 5=West Midlands; 6=East of England; 7=London; 8=South East; 9=South West |
| **4** | Ethnicity | 1 year before cohort entry | Cohort entry date | 1=White; 2=Non-white; 3=Not recorded |
| **5** | IMD quintile | 1 year before cohort entry | Cohort entry date | 1=Least deprived; 2=second least deprived; 3= middle deprivation; 4 second most deprived; 5=Most deprived |
| **6** | Comorbid stroke | 1 year before b cohort entry | Cohort entry date | 0=No; 1=Yes |
| **7** | Chronic kidney disease | 1 year before cohort entry | Cohort entry date | 0=No; 1=Yes |
| **8** | Hypertension | 1 year before cohort entry | Cohort entry date | 0=No; 1=Yes |
| **9** | Brain injury | 1 year before cohort entry | Cohort entry date | 0=No; 1=Yes |
| **10** | Heart disease | 1 year before cohort entry | Cohort entry date | 0=No; 1=Yes |
| **11** | Diabetic microvascular complications | 1 year before cohort entry | Cohort entry date | 0=No; 1=Yes |
| **12** | Peripheral vascular disease | 1 year before cohort entry | Cohort entry date | 0=No; 1=Yes |
| **13** | Anticoagulants | 6 months before cohort entry | Cohort entry date | 0=No; 1=Yes |
| **14** | Antiplatelet | 6 months before cohort entry | Cohort entry date | 0=No; 1=Yes |
| **15** | Statins | 6 months before cohort entry | Cohort entry date | 0=No; 1=Yes |
| **16** | Anticholinergics | 6 months before cohort entry | Cohort entry date | 0=No; 1=Yes |
| **17** | Antidepressants | 6 months before cohort entry | Cohort entry date | 0=No; 1=Yes |
| **18** | Antipsychotics | 6 months before cohort entry | Cohort entry date | 0=No; 1=Yes |
| **19** | Antihypertensives | 6 months before cohort entry | Cohort entry date | 0=No; 1=Yes |
| **20** | Glycated haemoglobin | 6 months before cohort entry | Cohort entry date | mmol/mol |
| **21** | Systolic blood pressure | 6 months before baseline | Cohort entry date | mmHg |
| **22** | Body mass index | 1 year before b cohort entry | Cohort entry date | $Kg/m^2$ |
| **23** | Smoking status | 1 year before cohort entry | Cohort entry date | 0=Non-smoker; 1=Current-smoker; 2=Ex-smoker |

†Cohort entry date is the date of the first initiation of a second-line antidiabetic medications of interest, also known as the index date, but does not mark the start of follow up.

**Table S4 Standardised mean difference (SMD) of each covariate before and after overlap weighting was applied**

| Covariate | Unweighted SMD[†] | Weighted SMD[†] |
|---|---|---|
| Sex | 0.106 | 0.011 |
| Smoking | 0.022 | 0.006 |
| Age at cohort entry | 0.338 | 0.036 |
| Body mass index | 0.293 | 0.014 |
| IMD quintile | 0.019 | 0.020 |
| Region | 0.005 | 0.030 |
| Ethnicity | 0.007 | 0.014 |
| Anticholinergic | 0.070 | 0.003 |
| Anticoagulants | 0.028 | 0.010 |
| Antidepressants | 0.065 | 0.009 |
| Antihypertensives | 0.051 | 0.030 |
| Antiplatelets | 0.151 | 0.049 |
| Antipsychotic | 0.031 | 0.006 |
| Statins | 0.057 | 0.040 |
| Chronic kidney disease | 0.071 | 0.026 |
| Hypertension | 0.006 | 0.034 |
| Systolic blood pressure (SBP) | 0.022 | 0.090 |
| Glycated haemoglobin (HbA1c) | 0.015 | 0.049 |
| Brain injury | 0.052 | 0.003 |
| Heart disease | 0.134 | 0.004 |
| Diabetic microvascular complications | 0.029 | 0.033 |
| Peripheral vascular disease | 0.023 | 0.004 |
| Comorbid stroke | 0.054 | 0.001 |
| Age at cohort entry : Body mass index | 0.336 | 0.035 |
| Systolic blood pressure : Glycated haemoglobin | 0.022 | 0.091 |

[†]The standardized mean difference (SMD) presented is the highest value of the levels of each covariate and their interactions, including the continuous variables that was modelled with 5 knots restricted cubic splines. The SMD of all the covariates were within the 0.10 threshold after applying overlap weighting method.

**Table S5 Cumulative Incidence rate of dementia per 100,000 person-years at risk**

| Outcome | TZD (n = 8,669) | DPP4 (n = 30,216) | SGLT2 (n = 29,429) | SU (n = 55,997) |
|---|---|---|---|---|
| Vascular dementia | 169 | 93 | 11 | 230 |
| All-cause dementia | 492 | 321 | 51 | 612 |

Abbreviations: TZD = thiazolidinedione; DPP-4 = dipeptidyl peptidase-4 inhibitors; SGLT2 = sodium–glucose cotransporter-2 inhibitors; SU = sulfonylurea.

**Table S6 Pairwise comparisons of incident dementia risks between second-line antidiabetic medications after accounting for death as a competing event**

| | Vascular dementia | | All-cause dementia | |
|---|---|---|---|---|
| **Comparison** | **aHR** | **95% CI** | **aHR** | **95% CI** |
| TZD – DPP-4 | 1.18 | (0.80–1.69) | 1.13 | (0.91–1.40) |
| TZD – SU | 0.76 | (0.57–1.02) | 0.88 | (0.70–1.04) |
| TZD – SGLT2 | 3.70 | (1.56–9.09) | 2.74 | (1.79–4.17) |
| SGLT2 – DDP-4 | 0.31 | (0.13–0.71) | 0.41 | (0.28–0.61) |
| SGLT2 – SU | 0.20 | (0.09–0.47) | 0.32 | (0.22–0.48) |
| DPP-4 – SU | 0.65 | (0.50–0.85) | 0.78 | (0.67–0.90) |

Adjustment for multiple comparisons was done using Tukey's method. Models were adjusted for age at baseline, sex, region, ethnicity, Index of Multiple Deprivation, comorbid stroke, chronic kidney disease, hypertension, heart disease, diabetic microvascular complications, peripheral vascular disease, concurrent use of anticoagulants, antiplatelets, statins, antihypertensives, anticholinergics, antidepressants, and antipsychotics, as well as biomarkers (systolic blood pressure, HbA1c) and lifestyle (body mass index, smoking status). Abbreviations: aHR = hazard ratio; CI = confidence interval; TZD = thiazolidinedione; DPP-4 = dipeptidyl peptidase-4 inhibitor; SGLT2 = sodium–glucose cotransporter-2 inhibitor; SU = sulfonylurea.

**Table S7 Pairwise comparisons of incident dementia risks between second-line antidiabetic medications with alternative definition of treatment initiation† (N = 88,427)**

| | Vascular dementia | | All-cause dementia | |
|---|---|---|---|---|
| **Comparison** | **aHR** | **95% CI** | **aHR** | **95% CI** |
| TZD – DPP-4 | 1.37 | (0.54–3.44) | 0.94 | (0.52–1.69) |
| TZD – SU | 0.95 | (0.39–2.33) | 0.75 | (0.42–1.28) |
| TZD – SGLT2 | 3.45 | (0.93–12.5) | 2.86 | (1.27–6.67) |
| SGLT2 – DDP-4 | 0.40 | (0.13–1.28) | 0.33 | (0.17–0.63) |
| SGLT2 – SU | 0.28 | (0.09–0.87) | 0.32 | (0.14–0.50) |
| DPP-4 – SU | 0.69 | (0.46–1.04) | 0.79 | (0.64–0.99) |

†Treatment initiation was redefined as either ≥2 prescriptions of the same SAM within 90 days, observed twice in 6-month intervals, or ≥2 prescriptions within one year, with gaps >90 and <180 days.

Adjustment for multiple comparisons was done using Tukey's method. Models were adjusted for age at baseline, sex, region, ethnicity, Index of Multiple Deprivation, comorbid stroke, chronic kidney disease, hypertension, heart disease, diabetic microvascular complications, peripheral vascular disease, concurrent use of anticoagulants, antiplatelets, statins, antihypertensives, anticholinergics, antidepressants, and antipsychotics, as well as biomarkers (systolic blood pressure, HbA1c) and lifestyle (body mass index, smoking status). Abbreviations: aHR = adjusted hazard ratio; CI = confidence interval; TZD = thiazolidinedione; DPP-4 = dipeptidyl peptidase-4 inhibitor; SGLT2 = sodium–glucose cotransporter-2 inhibitor; SU = sulfonylurea.

**Table S8 Pairwise comparisons of incident dementia risks between second-line antidiabetic medications using complete case analysis (N=99,240) and overlap weighting**

| | Vascular dementia | | All-cause dementia | |
|---|---|---|---|---|
| **Comparison** | **aHR** | **95% CI** | **aHR** | **95% CI** |
| TZD – DPP-4 | 1.20 | (0.70–2.04) | 0.86 | (0.37–2.00) |
| TZD – SU | 0.73 | (0.49–1.11) | 0.59 | (0.25–1.33) |
| TZD – SGLT2 | 2.94 | (0.93–9.09) | 1.54 | (0.45–5.26) |
| SGLT2 – DDP-4 | 0.41 | (0.14–1.20) | 0.56 | (0.21–1.46) |
| SGLT2 – SU | 0.25 | (0.09–0.74) | 0.38 | (0.15–0.99) |
| DPP-4 – SU | 0.61 | (0.43–0.88) | 0.68 | (0.52–0.89) |

Adjustment for multiple comparisons was done using Tukey's method. Models were adjusted for age at baseline, sex, region, ethnicity, Index of Multiple Deprivation, comorbid stroke, chronic kidney disease, hypertension, heart disease, diabetic microvascular complications, peripheral vascular disease, concurrent use of anticoagulants, antiplatelets, statins, antihypertensives, anticholinergics, antidepressants, and antipsychotics, as well as biomarkers (systolic blood pressure, HbA1c) and lifestyle (body mass index, smoking status). Abbreviations: aHR = adjusted hazard ratio; CI = confidence interval; TZD = thiazolidinedione; DPP-4 = dipeptidyl peptidase-4 inhibitor; SGLT2 = sodium–glucose cotransporter-2 inhibitor; SU = sulfonylurea.

**Table S9 Pairwise comparisons of incident dementia risks between second-line antidiabetic medications with a long follow-up period†**

| | Vascular dementia | | All-cause dementia | |
|---|---|---|---|---|
| Comparison | aHR | 95% CI | aHR | 95% CI |
| TZD – DPP-4 | 0.76 | (0.30–1.92) | 0.87 | (0.48–1.56) |
| TZD – SU | 0.51 | (0.21–1.27) | 0.61 | (0.34–1.09) |
| TZD – SGLT2 | 3.03 | (0.89–10.0) | 1.67 | (0.70–4.00) |
| SGLT2 – DDP-4 | 0.25 | (0.10–0.64) | 0.52 | (0.26–1.01) |
| SGLT2 – SU | 0.17 | (0.07–0.42) | 0.36 | (0.19–0.71) |
| DPP-4 – SU | 0.68 | (0.47–0.97) | 0.70 | (0.57–0.86) |

†Follow-up started from 180 days after the first second-line antidiabetic medication prescription and ended on the transfer out of practice, loss to follow-up, dies or the study ends (31/12/2023), whichever occurs first.

Adjustment for multiple comparisons was done using Tukey's method. Models were adjusted for age at baseline, sex, region, ethnicity, Index of Multiple Deprivation, comorbid stroke, chronic kidney disease, hypertension, heart disease, diabetic microvascular complications, peripheral vascular disease, concurrent use of anticoagulants, antiplatelets, statins, antihypertensives, anticholinergics, antidepressants, and antipsychotics, as well as biomarkers (systolic blood pressure, HbA1c) and lifestyle (body mass index, smoking status). Abbreviations: aHR = hazard ratio; CI = confidence interval; TZD = thiazolidinedione; DPP-4 = dipeptidyl peptidase-4 inhibitor; SGLT2 = sodium–glucose cotransporter-2 inhibitor; SU = sulfonylurea.

**Figure S1 Distributions of generalized propensity scores stratified by treatment groups, and showing poor overlap†**

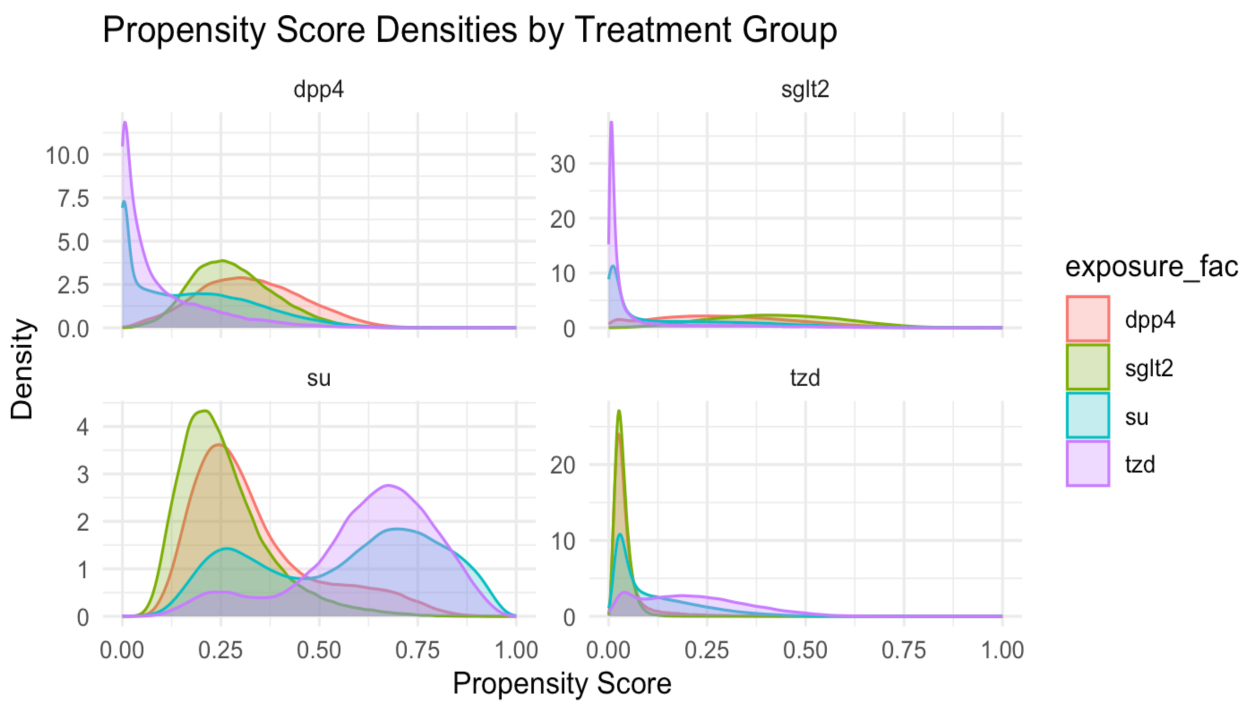

†The generalized propensity scores were defined as the probability of receiving each specific second-line antidiabetic medication, conditional of the observed patient baseline characteristics, estimated using multinomial logistic model.
Abbreviations: TZD = thiazolidinedione; DPP-4 = dipeptidyl peptidase-4 inhibitor; SGLT2 = sodium–glucose cotransporter-2 inhibitor; SU = sulfonylurea.

**Figure S2 Post-weighting standardised mean difference (SMD) using inverse probability treatment weighting (IPTW)**

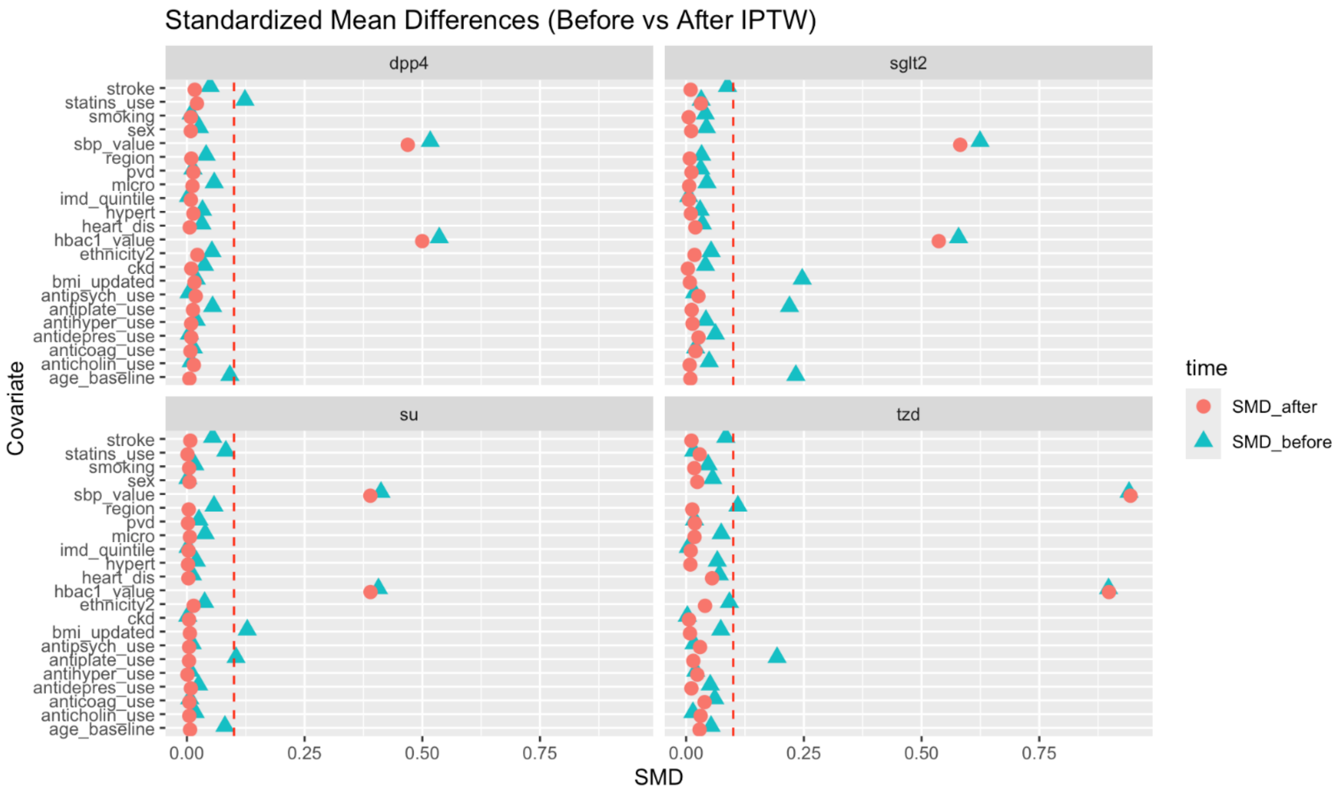

Note: Before IPTW weighting the SMD of some covariates were > 0.1 threshold, after IPTW was applied the SMD of most covariates was contained within the threshold, however the SMDs of SPB and HbA1c were still high (> 0.30). Time implies the period before and after the application of IPTW. The propensity score model included age at baseline, sex, region, ethnicity, Index of Multiple Deprivation, comorbid stroke, chronic kidney disease, hypertension, heart disease, diabetic microvascular complications, peripheral vascular disease, concurrent use of anticoagulants, antiplatelets, statins, antihypertensives, anticholinergics, antidepressants, and antipsychotics, as well as biomarkers (systolic blood pressure, HbA1c) and lifestyle (body mass index, smoking status). Abbreviations: TZD = thiazolidinedione; DPP-4 = dipeptidyl peptidase-4 inhibitor; SGLT2 = sodium–glucose cotransporter-2 inhibitor; SU = sulfonylurea.

### File S1 Covariates generated with published algorithms

#### Smoking status

Patients' smoking status were derived as 5 categories: non-smokers, ex-smokers, moderate smokers, light smokers and heavy smokers based on the most recent number of cigarettes a patient used to smoke in a day (1). We re-classified the 5 groups to three: non-smokers, current-smokers (a combination of moderate, light and heavy smokers), ex-smokers to improve sample size in each group ensuring model convergence. Where the most recent record of a patient is a non-smoker, but previous records indicate a history of smoking, the smoking status for such an individual is updated to ex-smoker. For more details on the algorithm used to create the smoking status variable please visit the R vignette of CPRD package (https://alexpate30.github.io/rcprd/articles/Details-on-algorithms-for-extracting-specific-variables.html) (1,2).

#### Systolic Blood Pressure

We used the most recent Systolic Blood Pressure (SBP) record measured within one year before baseline according to National Institute for Health and Care Excellence (NICE) guidance. NICE recommends measuring blood pressure at least once in a year, for those with T2DM (3). SBP values <40 mmHg or >270 mmHg were excluded. For individuals with multiple measurements on the same date, the daily mean SBP was used. The standard unit of measurement was millimetres of mercury (mmHg); records with other or unknown units were excluded.

#### Glycated haemoglobin - HbA1c

NICE recommends that HbA1c be measured within 3 – 6 months in people with T2DM (4); therefore, we used HbA1c values recorded within the 6 months preceding SAM initiation. HbA1c was recorded in mmol/mol; records with other or unknown units were excluded. Pre-2011 values reported in Diabetes Control and Complications Trial (DCCT) percentage units were converted to mmol/mol using the published formula (%value − 2.15) × 10.92. Values <10 mmol/mol or >200 mmol/mol were excluded (5). For those with multiple measurements on the same date, daily mean HbA1c was.
